# Uncovering the fitness of endemically circulating Zika virus strains

**DOI:** 10.64898/2026.06.21.26356168

**Authors:** Yining Chen, Douglas Fritz, Hannah Clapham, Noemie Lefrancq, Henrik Salje, the BUZZ study team

**Author notes:** Corresponding author: Henrik Salje. <u>BUZZ study team (listed in alphabetical order):</u> Pablo Aguilar Ticona, Kathryn Anderson, Darunee Buddhari, Derek Cummings, Marco Hamins-Puertolas, Taweewun Hunsawong, Ricardo Khouri, Albert Ko, Kathryn McGuckin, Nivison Nery Jr., Mitermayer Reis, Stephen J. Thomas, Adam T. Waickman.

## Abstract

Zika virus (ZIKV) is an arbovirus that usually causes few symptoms and has circulated endemically in Asia for decades. However, a large outbreak in South America in 2015 uncovered the serious risk of congenital Zika syndrome in infants born from ZIKV infected mothers. It is unknown whether a lineage with distinct pre-existing fitness advantage emerged from Asia to cause the South American outbreak, and whether there is ongoing evolution that can result in future globally fit strains. Here we used 107 sequences from a single setting (Thailand) collected over an 18 year period (2006-2023). We used novel analytical tools to identify distinct lineages that have circulated in the population and estimated their relative epidemiological fitness. We found there have been six lineages circulating sequentially in the country, with regular emergence and replacement of lineages showing higher fitness than their predecessors. We identified 15 lineage-defining amino acid changes, including four well-documented fitness-enhancing mutations, and two UTR substitutions. The lineage that emerged in South America was evolutionarily linked to the highest-fitness lineage in Thailand, carrying seven of our lineage-defining substitutions acquired during endemic circulation there, and subsequently accumulating four additional changes. After the global pandemic, endemic ZIKV in Thailand continued to evolve, with newly emerged lineages showing novel mutations and increased fitness. Our findings have key implications for the monitoring of ZIKV and can help identify the pathway to increased transmissibility of this globally important pathogen.

## Introduction

Zika virus (ZIKV) is an arbovirus within the *Flavivirus* genus, spread by *Aedes albopictus* and *Aedes aegypti* mosquitoes. There are two major lineages of ZIKV. The African lineage, first discovered in Uganda in 1947 and the Asian lineage, first identified in Malaysia in 1966, which has circulated endemically in Southeast Asia for decades *(1)*. ZIKV typically causes subclinical infection or a mild, self-limiting febrile illness, frequently presenting with malaise, rash, and anorexia *(2)*. This subtle clinical profile has contributed to its long-term silent circulation and the historical lack of large-scale detection. In 2013, a large outbreak caused by the Asian lineage was observed in French Polynesia. In 2015, the Asian lineage spread across the Pacific to the Americas, culminating in an unprecedented epidemic, particularly in Brazil. ZIKV infections during these large outbreaks revealed a strong association between infection in pregnant women and congenital Zika syndrome (CZS) in their infants *(3)*. The epidemiological discrepancy between the devastating epidemics in the Americas and the historically low-level endemic circulation in Southeast Asia underscores a pivotal uncertainty regarding the evolutionary origins of the ZIKV strains that emerged in the Americas. It also remains unclear whether the Asian-lineage ZIKV strains circulating in Southeast Asia possess epidemic potential comparable to the variants that drove the outbreaks in the Americas.

Characterizing viral evolutionary trajectories and fitness dynamics across shifts in transmission efficiency aids in deciphering drivers of emergence. Phylogenetic analysis of the epidemic lineage in the Americas identified several key amino acid substitutions *(4, 5)*. Yet, the evolutionary precursors of the South American virus are rooted in Southeast Asia, which represents the primary genomic reservoir of the Asian ZIKV lineage. Within this region, decades of low-level circulation have generated extensive viral genetic diversity. Elucidating the evolution and fitness variation of ZIKV in Southeast Asia aids in understanding whether epidemiologically fitter variants ultimately emerged to cause global pandemics. However, the evolutionary dynamics of ZIKV in Southeast Asia remain largely uncharacterized. Owing to low disease severity and clinical similarity to other co-circulating flaviviruses, the virus was under-reported within this region, leaving significant gaps in our understanding of its diversification and fitness.

In this study we used ZIKV genome sequences collected since 2006 from Thailand, a key endemic hub in Southeast Asia, to investigate the viral population dynamics preceding and following the major phase of global pandemic. Critically, there exists a large number of ZIKV sequences available from this single location over a long time period, which therefore provides an opportunity to understand whether there are distinct lineages that have co-circulated in a single ecological setting. We used a novel framework to identify lineages based on viral fitness and phylogenetic topology *(6)*. Our analysis reveals the emergence of sublineages with increasing fitness in Southeast Asia, occurring in parallel with the rise of the variants that spread to French Polynesia and Brazil. The genomic divergence identified between these Southeast Asian sublineages and the epidemic strains provides a deeper understanding of ZIKV fitness landscape and helps contextualize future emergence risk.

## Results

### Genetic diversity of ZIKV in Thailand

We built a time-resolved phylogeny of 107 sequences from Thailand during 2006-2023 and 80 sequences from Brazil during 2015-2019 (Figure 1a-b). The Thai sequences showed substantial genetic diversity consistent with long-term ZIKV circulation. We estimated from these sequences that ZIKV emerged in Thailand in February 2000 (95% Highest Posterior Density (HPD) intervals: October 1998-April 2001). The Brazilian sequences formed a well-supported monophyletic clade (Posterior Probability (PP): 0.95) nested within the diversity of the Thai sequences.

**Figure 1.**
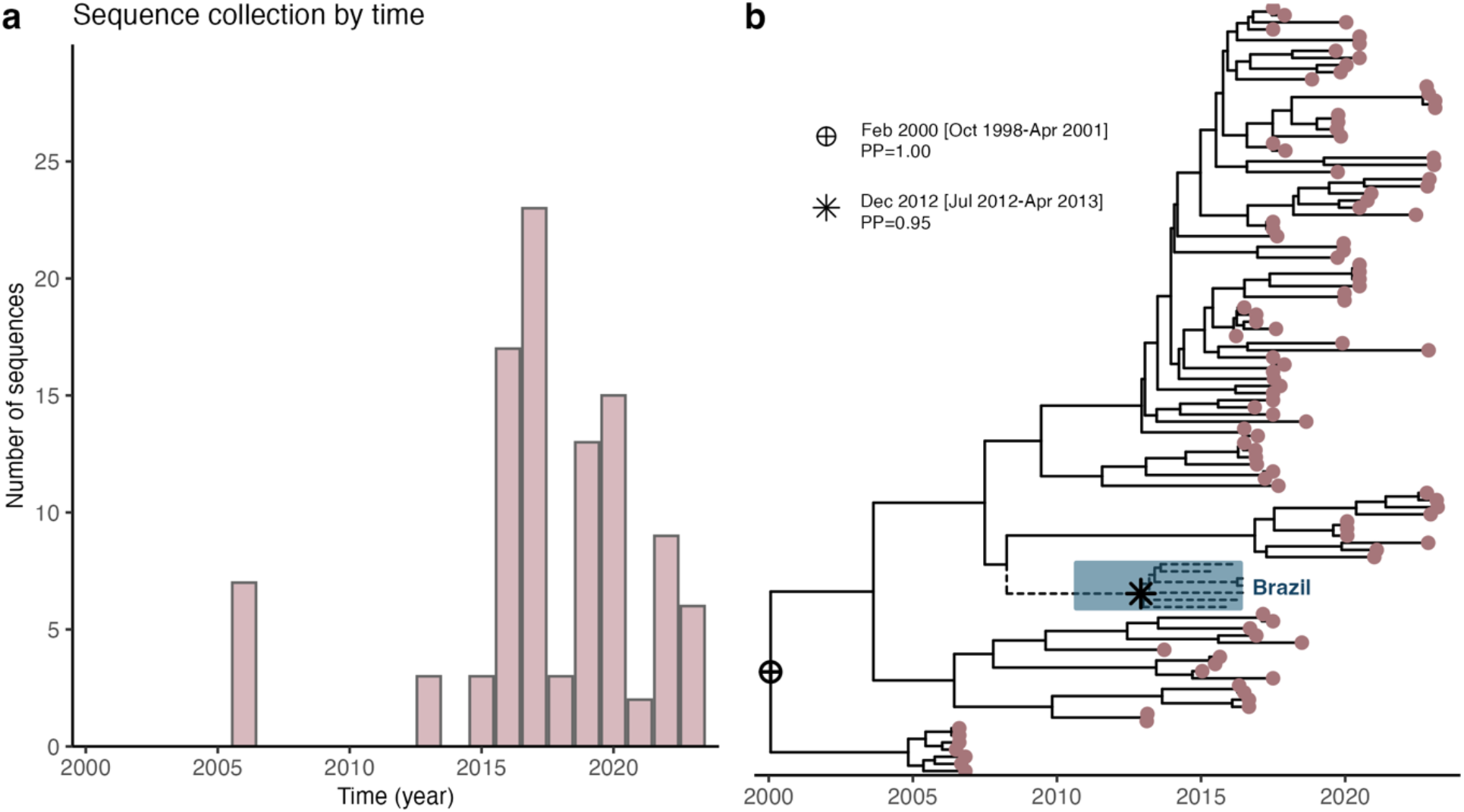
Temporal distribution and phylogenetic of ZIKV sequences in Thailand. a. Annual number of ZIKV sequences collected in Thailand by calendar year. b. The time-resolved maximum clade credibility phylogenetic tree for the Thailand sequences (coloured tips) and Brazil sequences (dashed branches; shaded background). Selected internal nodes are marked with symbols and annotated as estimated node date [95% Highest Posterior Density (HPD)], Posterior Probability (PP).

We adapted a phylogenetic clustering model to use the phylogeny to agnostically cluster the Thai sequences into distinct lineages based on distinct levels of fitness *(6)*. This approach relies on epidemiologically fitter viruses having more closely genetically related descendants than other viruses circulating at the same time. Using this approach, we identified six distinct lineages (termed Groups 1-6) that circulated in Thailand between 2000 and 2023 (Figure 2a-b). We estimated that the Brazilian strains were evolutionarily related to Group 4, sharing a closer common ancestor than other lineages (PP: 0.77). However, there was a long branch (8.60 years) from this common ancestor to Group 4, compared with the branch (4.66 years) leading to the Brazilian strains, consistent with additional unsampled diversity around this split prior to emergence in Brazil.

**Figure 2.**
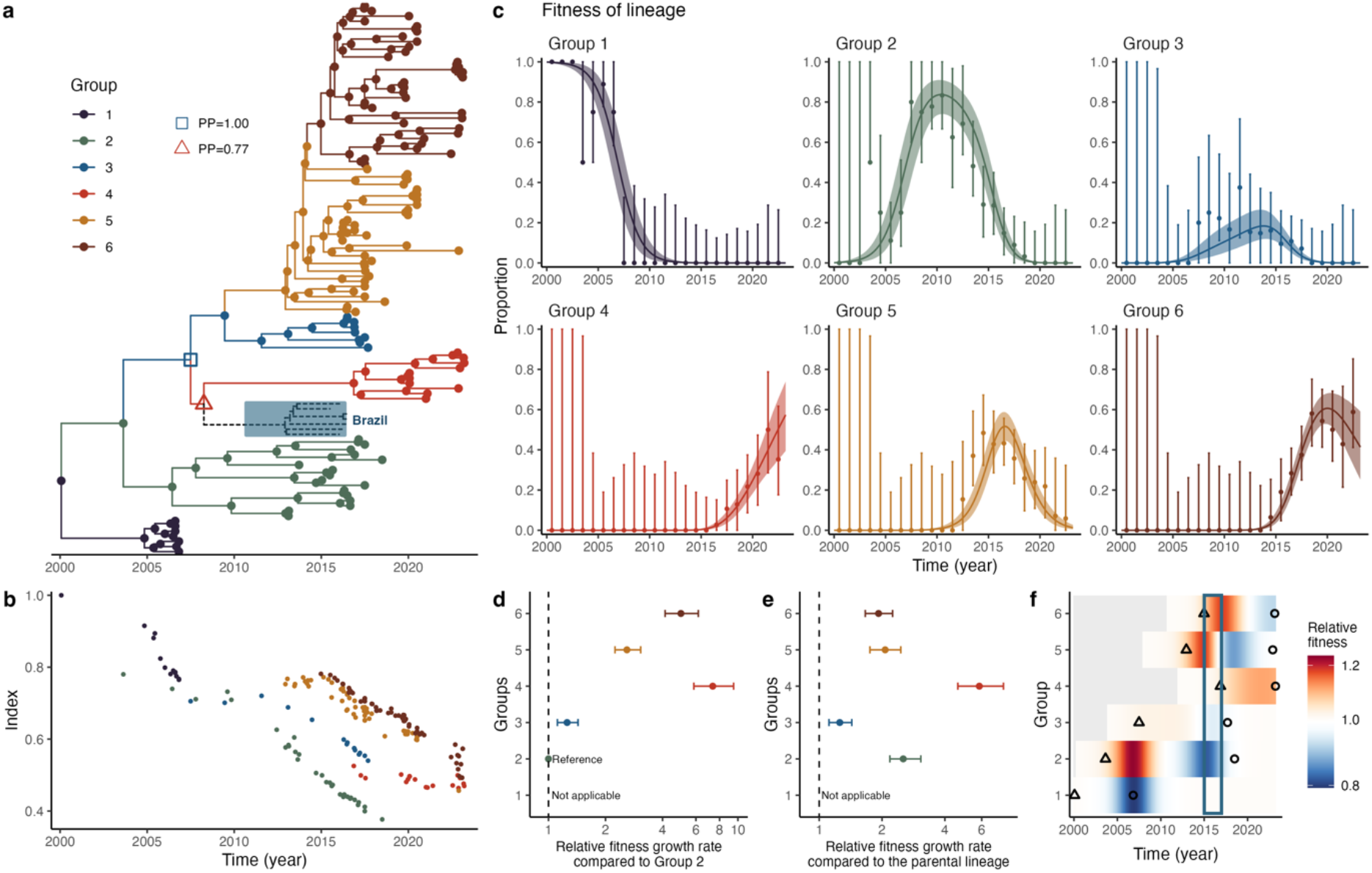
Identified ZIKV lineages and their fitness estimates. a. Identified ZIKV lineages on the time-resolved phylogeny. Brazilian sequences (dashed branches; shaded background) were presented for phylogenetic context and not included in the lineage identification analysis. a. Abbreviations: PP, Posterior Probability. b. The index dynamics for internal and external nodes, computed as a kernel-weighted genetic-distance measure to capture lineage emergence (see Materials and Methods for details). c. The fitness dynamics of the identified lineages. The points and vertical bars refer to empirical values and 95% confidence intervals derived from the phylogenetic tree. The solid lines and shaded regions refer to the posterior median and 95% credible intervals estimated from the fitness model (see Materials and Methods for details). d-e. The estimates of relative fitness growth rate compared to the ancestral Group 2 (d) and the parental lineage (e). The points and bars indicate posterior median and 95% credible intervals. Estimates relative to the oldest lineage, Group 1, are shown in Supplementary Fig. S1. f. The real-time relative fitness compared to the contemporary co-circulating lineages. Triangles indicate the earliest internal node within each lineage, and points indicate the latest sampled tip within that lineage. The vertical box indicates the timeframe of the initial large-scale ZIKV outbreak in Brazil (2015–2016).

### Relative fitness of each lineage

We identified a clear succession of ZIKV lineages over the 24 years in Thailand (Figure 2c). Prior to 2006, the viral population was dominated by Group 1. Following 2007, Group 2 achieved epidemiological prominence, maintaining a high population frequency until 2012 and persisting in circulation until 2018, alongside the moderate co-circulation of Group 3. Beginning around 2012, Group 5 underwent rapid expansion, eventually reaching co-dominance with Group 2. From approximately 2015, Groups 4 and 6, following their earlier diversification, entered a phase of sustained expansion.

We fitted a multinomial logistic regression model to the number of nodes and tips in the tree that belonged to each lineage in each year, assuming a constant growth rate for each lineage. The estimates were similar when based on tip counts only (Supplementary Fig. S1). This allowed us to quantify the relative fitness of each lineage. This simple model was able to recover the observed frequencies of each lineage per year (Figure 2c). We estimated that Group 3 viruses had 1.25 (95% confidence intervals (CI): 1.11-1.43) times the fitness of Group 2 viruses (our reference group), where fitness is defined as the relative growth of a virus per year (Figure 2d). By contrast, Groups 4-6 had much higher levels of fitness, with relative growth ranging between 3-7 relative to Group 2. We found Group 4 had the greatest fitness advantage, with a 7.4 (95% CI: 5.9-9.5) relative fitness as compared to Group 2 and 5.9 (95% CI: 4.6-7.6) as compared to its parent lineage (Group 3) (Figure 2d-e). Group 6 also had a high relative fitness, with a 5.0 (95% CI: 4.1-6.2) relative fitness as compared to Group 2, despite a less pronounced fitness advantage of 1.9 (95% CI: 1.7-2.3) compared to its parent lineage (Group 5) (Figure 2d-e).

We next estimated the changing relative fitness of each lineage, relative to what else was circulating at the time (Figure 2f). This allows us to consider that the success of a lineage depends on the fitness of all other strains present at the same moment in time, ignoring extinct (and yet to emerge) lineages. We found that there was a chronological succession in fitness dominance: Group 2 exhibited the primary growth advantage during 2005–2008, followed by Group 3 (2009–2012), Group 5 (2013–2015), Group 6 (2016–2018), and Group 4 (2019–2023). There was an average of 3 lineages circulating within any year (range 2005-2023), with the mean lifespan of each lineage of 9 years.

### Lineage-defining substitutions

We compared the sequences in each of the lineages to identify lineage-specific amino acids. Overall, we identified 17 lineage-defining substitutions, including 15 amino acid substitutions and 2 untranslated region (UTR) substitutions (Figure 3). The amino acid changes were distributed throughout the genome with three in each of NS2A and NS2B, two in E, prM, NS4B, NS5, NS1 and a single change in C. There were also two changes in the 3’ UTR, located upstream of the SL1 structural domain (position 7) and between the DB1 and DB2 structural domains (position 234), respectively (Figure 3). We found that seven of these changes occurred between Group 1 and Group 2. For subsequent Groups, there were only between one and three amino acid changes. The exception was Group 4, where there were six changes. We note the long branch linking group 4 with its parent lineage, suggesting a possible novel introduction into the country. The lineage which emerged in Brazil was identical to Group 5 at all of these 17 sites. In addition, however, the Brazilian lineage possessed four extra changes, including one in each of prM and NS5 and two in the 3’ UTR (located upstream of SL1 and proximal to SL2 respectively). The lineage that emerged in French Polynesia, prior to Brazil, was also identical to Group 5 at these 17 lineage-defining sites and 2 of the extra positions, while sharing the remaining two changes ultimately seen in Brazil.

**Figure 3.**
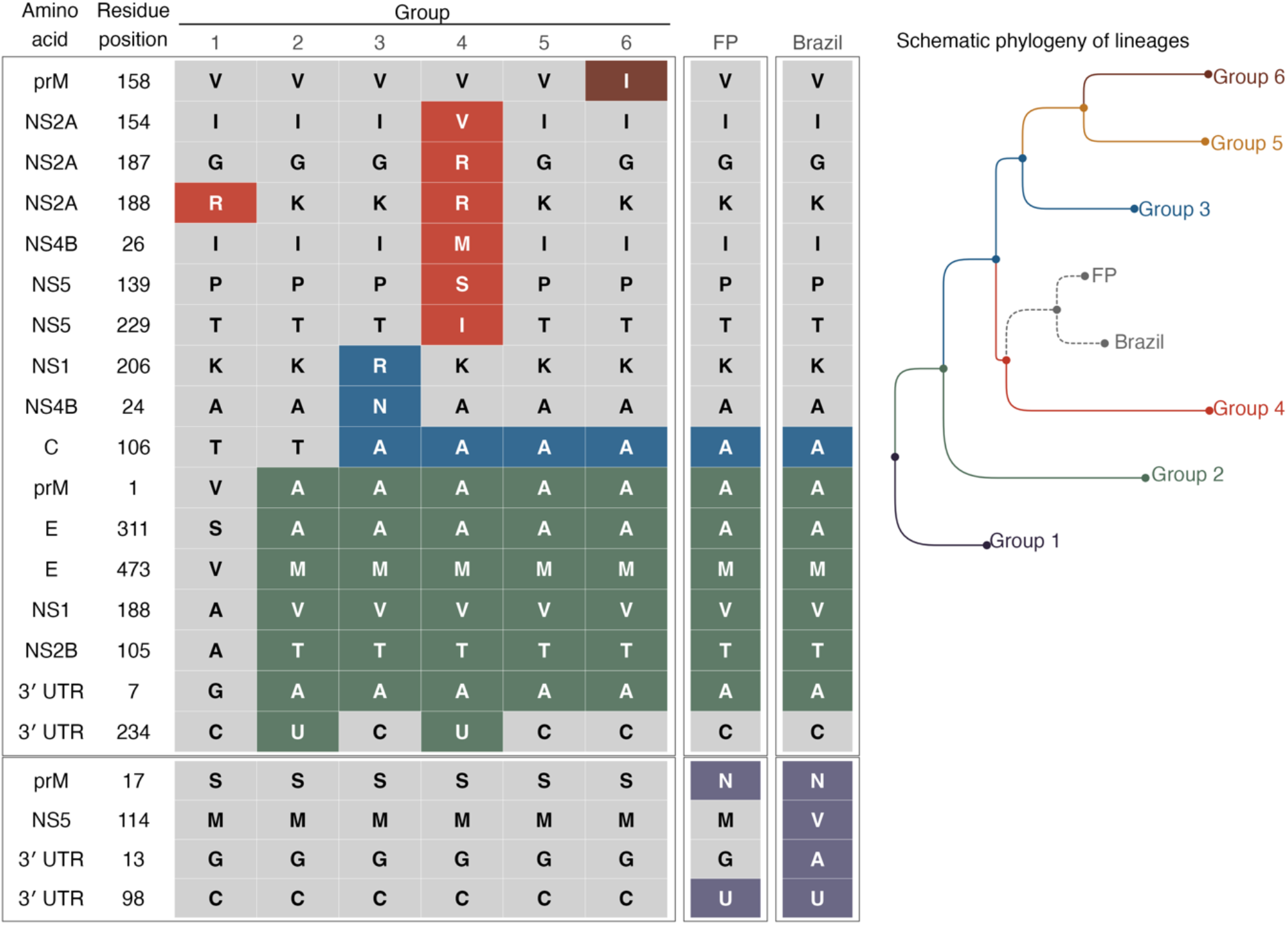
Lineage-defining substitutions: Amino acid substitutions and 3′ untranslated region (UTR) nucleotide substitutions. Rows correspond to specific residue positions, with letters representing the consensus residue for each lineage. Nucleotide positions of 3′ UTR are numbered according to the ZIKV reference sequence KX197192. Shaded cells indicate the presence of substitutions, with colours indicating the lineage defining each substitution. The distribution of these substitutions on the phylogenetic tree is detailed in Supplementary Figs. S2-S3. Abbreviations: FP, French Polynesia.

## Discussion

Here we have used sequences from a single endemic ecological setting to investigate the ecology of ZIKV. We found that there are multiple ZIKV lineages highly dynamic, with clear evidence indicating lineages emerge with distinct fitness advantages over previously circulating lineages. The strain that emerged in Brazil had a very similar amino acid sequence to the strains present in Thailand at the time. However, since the global outbreak, endemic ZIKV has continued to evolve in Thailand with the emergence of novel strains showing further increased fitness relative to the local prior lineages in the populations.

It remains unclear why ZIKV caused a single major outbreak in South America while it continues to circulate endemically in Asia. We found that while the ZIKV lineage that emerged from Asia to South America did have some of the key fitness advantages, it also acquired at least two extra amino acid changes - either in South America or in an intermediary location. Further, while there have been ZIKV reintroductions from South America back to Asia, we have not observed a clear displacement of the indigenous strains, with locally circulating ZIKV lineages in Thailand continuing to acquire additional fitness advantages.

Beyond intrinsic viral characteristics, host population heterogeneities may play a substantial role in shaping both evolutionary dynamics and epidemiological outcomes. Our endemic lineage Group 4 shared a closer recent common ancestor with the Brazil strain compared to other lineages. The viral evolutionary trajectories diverged when circulating in differing host landscapes. Brazil strain evolves in a highly susceptible host population. Group 4 evolves in an endemic region where the host population has a basic level of immunity, and is genetically distinct from the other lineage. Meanwhile, high host susceptibility creates an inherent vulnerability to large-scale outbreaks, regardless of viral intrinsic fitness. A similar rapid expansion across South America following its introduction was observed with Chikungunya in 2014 *(7)*.

The immune interaction between dengue virus (DENV) and ZIKV is complex. Across both South America and Southeast Asia there has been widespread DENV circulation for decades leading to substantial population immunity. Despite regionally synchronous epidemics, DENV viral diversity is spatially constrained with limited DENV flow from Thailand to neighbouring countries *(8, 9)*. This means that DENV evolution may be setting specific. The regional DENV immunological landscapes and viral profiles potentially modulate ZIKV viral divergence and disease epidemiology. Future studies investigating the role of these immune interactions are warranted to better understand the setting-specific ZIKV evolution and disease dynamics.

Some of our identified amino acid substitutions are well documented in the literature. The residues prM-17, NS1-188, and NS5-114 have been used as primary markers for classifying Asian-lineage ZIKV genotypes *(4)*. Notably, the Southeast Asian endemic lineages lacked both the prM-S17N or NS5-M114V substitutions (Figure 3). These substitutions show varying functional impacts on ZIKV viral fitness and pathogenesis. The NS1-A188V substitution, which was present in our early-circulating Group 2 and remains fixed, has been reported to enhance vector adaptation and facilitate viral replication in humans by antagonizing interferon-β (IFNβ) production *(10)*. The prM-S17N substitution has been associated with increased infectivity in neural progenitor cells and exacerbation of microcephaly phenotypes *(11)*, underscoring its potential contribution to the heightened severity of the ZIKV epidemic in South America. Nevertheless, the extent to which these pathogenic effects translate into enhanced transmission efficiency or overall viral fitness remains unclear. While NS5-M114V was highly specific to the American strain, some studies suggested that it was unlikely to be a primary determinant of replication efficiency or transmission potential *(12)*.

In addition, our lineage-defining substitutions C-T106A, PrM-V1A, and E-V473M, present since the earlier lineages Groups 2-3, have been functionally documented in significantly enhancing viral infectivity and epidemic potential. C-T106A has been reported to promote efficient virion assembly and increase viral transmission potential *(13)*. PrM-V1A showed strong association with increased virulence and faster growth in mosquito cell lines. Experimental models suggested that it may elevate higher viremia and contribute to severe fetal outcomes, including early-stage fetal loss that may precede the development of microcephaly *(14)*. E-V473M has been shown to enhance virion morphogenesis and increase viremia in non-human primates, which was considered a critical determinant for enhanced intrauterine transmission and urban spread *(15)*.

In contrast to the well-documented markers, the substitutions E-S311A and NS2B-A105T, identified in our Groups 2-6 and in the French Polynesia and Brazilian strains, remain comparatively understudied. Residue 311 of the envelope protein, together with residues 310 and 333, forms part of a structural epitope in which single amino acid substitutions have been shown to mediate antibody escape *(16)*. The limited research focus on these sites may be related to their conservation across both historical Malaysian strains and contemporary epidemic lineages (Supplementary Fig. S4). Because this shared genetic state is maintained across major lineages, their potential evolutionary relevance may be overlooked in conventional comparative analyses and only becomes apparent when contrasted against a specific ancestral baseline, such as Group 1. Apart from the envelope residue 311, no other lineage-defining substitutions directly overlapped with reported ZIKV monoclonal antibody escape sites or candidate antiviral-resistance mutations.

Our analysis further identified dynamic shifts at residues NS1-206 and NS4B-24. These substitutions initially appeared in the intermediate lineage Group 3, but reverted to the ancestral state in subsequent descendant lineages. Notably, Group 5, which diverged from Group 3, exhibited these reversions alongside a significant increase in estimated viral fitness. Particularly, Group 6, which evolved from Group 5 and was associated with further fitness gains, was characterized by a novel lineage-defining substitution prM-V158I. The residue 158 is highly conserved across the ZIKV phylogeny that the ancestral valine (V) has remained stable across the other identified Southeast Asian endemic lineages, the American epidemic strains, as well as the earlier 1966 Malaysian and African lineages. The emergence of a stable isoleucine (I) at this position represents a notable evolutionary divergence that coincides with the high expansion potential of Group 6.

The contemporary lineage Group 4, which shows greatest fitness advantage, is characterized by six unique lineage-defining substitutions, reflecting its marked genetic divergence and long branch from the parental ancestor (Figure 3). The mutations NS2A-I154V, NS2A-G187R, NS2A-K188R, NS5-P139S, and NS5-T229I, were detected in the 2020–2023 Thailand cases and in the 2019 Myanmar strain *(17)*. Notably, the mutations NS4B-I26M, NS5-P139S, and NS5-T229I represent a reversion to the ancestral African or early Asian (1966) states (Supplementary Fig. S4). Although most documented ZIKV outbreaks were caused by the Asian lineage, the African lineage has been shown to be more pathogenic in both vitro and vivo models *(18, 19)*. Similar mutational reversions were observed at residues NS1-188, C-106, and prM-1, which have been shown to have an effect on fitness restoration in the literature *(5)*. In addition, NS4B-I26M has been reported to be enriched in infected mouse brain tissue and may act in concert with an NS1 variant, suggesting a potential contribution to neurotropism *(20)*. Although the functional impact of these mutations remains less characterized, their unique presence in this expanding lineage suggests a potential contribution to its inferred elevated fitness and warrants further experimental investigation.

A limitation of this study is the relatively small number of available sequences, particularly during the period preceding the global ZIKV outbreaks. Despite the long-term endemicity of ZIKV in Southeast Asia, surveillance has historically been limited by the virus’s typically mild clinical presentation and its symptomatic similarity to other co-circulating flaviviruses. This resulted in sparse data from 2006, a sampling gap between 2007 and 2012, with a gradual increase in sequence availability after the 2015 Brazil outbreak. This sampling gap accounts for the accumulated amino acid substitutions observed between the basal lineage Group 1 and the early circulating lineage Group 2. Nevertheless, the available sequences remain robust enough to reconstruct the endemic virus evolutionary trajectory, providing essential insights into the viral fitness and genetic shifts that occurred before and after the virus expanded globally.

In conclusion, this study characterizes the highly dynamic evolution of endemic ZIKV in Southeast Asia, identifying six distinct lineage transitions during 2000-2023. These lineages consistently exhibited increased fitness growth relative to their progenitors, driven by specific amino acid and UTR substitutions. Our findings suggest that ZIKV accrued several fitness-associated changes during this endemic period prior to its dissemination to South America. The subsequent global outbreaks likely resulted from a synergy between these pre-existing fitness advantages and further localized viral adaptation, amplified by high host susceptibility and complex immunological interactions with co-circulating flaviviruses. Notably, in parallel to the global epidemics, the endemic ZIKV continued to evolve along a divergent trajectory, yielding new lineages with elevated fitness and novel mutations. These results underscore the necessity for heightened surveillance of contemporary endemic lineages in Southeast Asia to better evaluate epidemic risk. Furthermore, our findings motivate future experimental work to characterize the impact of these emerging substitutions on viral phenotypes, clinical outcomes, and epidemic potential, providing critical insights for outbreak preparedness.

## Materials and Methods

### Sequence data

ZIKV nucleotide sequences collected in Thailand were retrieved from the National Centre for Biotechnology Information (NCBI) database, accessed in October 2025. This primary dataset included one sequence from a traveller to Thailand identified in Japan (accession number: LC369584.1). Sequences with a length < 1,000 bp were excluded. A total of 107 Thailand ZIKV sequences (collected 2006-2023) were used for the core phylogenetic analysis.

For contextual analysis, ZIKV sequences with a length > 10,000 bp from French Polynesia (*N* = 14) and Brazil (*N* = 80) were extracted from the NCBI database. These external sequences served exclusively as reference data for comparing the identified Thailand sublineages against known epidemic strains.

All sequences were aligned using Multiple Sequence Comparison by Log-Expectation (MUSCLE) in software MEGA (version 11) with the default settings *(21)*. The resulting alignment was manually checked to ensure the absence of frameshift mutations. The final alignment comprised 11,171 nucleotide positions.

### Phylogenetic reconstruction

The time-resolved phylogenetic tree was inferred using BEAST (version 10.5.0). We used a GTR substitution model, a strict molecular clock, and a constant-size coalescent prior. To improve parameter mixing, the Bayesian sampling weight for the clock rate was increased from 3 (default weight) to 30. Three independent Markov Chain Monte Carlo (MCMC) chains were run for 20,000,000 iterations each, with parameters sampled every 1,000 iterations. The first 10% samples from each run were removed as burn-in. The remaining samples were combined and thinned to a sampling frequency of every 2,000 iterations using LogCombiner (version 10.5.0). Convergence was assessed in Tracer (version 1.7.2) that effective sample sizes were > 200 for all parameters. A maximum clade credibility (MCC) tree with median node heights was generated using TreeAnnotator (version 10.5.0).

Analysis of lineage detection was performed on the subtree containing only Thailand ZIKV sequences. Six tips descending from nodes subtending branches longer than seven years were excluded (see Supplementary Fig. S5 for the excluded sequences). This avoided potential bias from temporally isolated clades that may not reflect ongoing transmission and could better capture co-circulating lineages within the endemic setting.

### Lineage detection using phylowave

We applied phylowave, a previously published algorithm for automated lineage identification from time-resolved phylogenetic trees *(6)*. Briefly, for each node *i* (internal and terminal) sampled at time *t*_*i*_, we computed an index *D*_*i*_ to summarize its relative genetic proximity to the rest of the population circulating at time *t*_*i*_. It was calculated as a kernel-weighted average of genetic distances from node *i* to all other contemporary nodes *j*, using an exponential kernel to weigh shorter genetic distances more heavily. For computational efficiency, *D*_*i*_ was calculated from pairwise distances *d*_*i,j*_ as

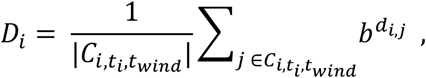

where time window *t*_*wind*_ is a prefixed parameter; 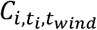 represents the set of other circulating nodes (*j* ≠ *i*) during the time interval [*t*_*i*_ − *t*_*wind*_, *t*_*i*_ + *t*_*wind*_], defined by the presence of branches traversing this period; *b* ∈ [0,1] is the bandwidth, calculated as the characteristic time to the most recent common ancestor, and is derived from a specified timescale, the ZIKV genome length, and the mutation rate; *d*_*i,j*_ denotes the pairwise genetic distance between nodes *i* and *j*, approximated as the cumulative number of substitutions between them on the time-resolved phylogeny.

Distinct viral populations are expected to produce lineage-specific temporal trajectories of the index, often manifesting as differentiated expansion patterns. To capture the time-varying fitness among the co-circulating sublineages, we modelled the log-transformed index dynamics using a generalized additive model (GAM) with penalized cubic splines. For each node *i* occurring at time *t*_*i*_, we expressed log*D*_*i*_ as a linear additive predictor comprising a global intercept, a baseline spline for time *t*_*i*_ with *k* knots (*B*_0_(*t*_*i*_, *k*)), and a lineage-specific spline for lineage *l* that *i* belongs to (*B*_*l*_(*t*_*i*_, *k*)). The algorithm begins with a single viral population, and the temporal variation in the index is captured solely by the baseline spline *B*_0_. At each subsequent iteration, one additional lineage is introduced by identifying the node whose partition best explains heterogeneity in index dynamics, as measured by the increase in deviance explained by the model. This iterative partitioning continues until adding an additional lineage fails to improve model performance beyond a predefined threshold.

In this study, we set the time-window parameter to *t*_*wind*_ = 2 years for index calculation. The timescale was set to 5 years for bandwidth calculation, using a genome length of 11,171 bp and a substitution rate of 6.8 × 10^−4^ per site per year, taken from the posterior estimate under the BEAST time-scaled phylogeny. The stopping criterion in lineage identification algorithm was defined as an increase in deviance explained of less than 0.5% at a given step. Candidate split nodes were restricted to those with at least 10 descendant tips, and the minimum lineage size was set to 10 nodes. Penalized cubic splines were fitted with *k* = 3 knots. To account for uneven temporal sampling, isolates were weighted according to sampling frequency within 0.5-year time windows in GAM. All analyses were implemented in R version 4.2.1 using the published phylowave codebase *(6)*.

### Viral fitness estimation

To quantify the fitness of the identified lineages, which was measured as the temporal dynamics of lineage proportions, we applied a Bayesian multinomial logistic model adapted from Lefrancq et al *(6)*. Let *L* denote the number of lineages, *y*_·,*t*_ = {*y*_*l,t*_: *l* = 1, · · ·, *L*} denote the vector of isolate counts for each lineage *l* at time *t* . We assumed that *y*_·,*t*_ follows a multinomial distribution parameterized by the expected lineage proportions *θ*_·,*t*_ = {*θ*_*l,t*_ ∈ [0,1]: *l* = 1, · · ·, *L*}, where *Σ*_*l*_*θ*_*l,t*_ = 1. *θ*_·,*t*_ was modelled using a softmax function:

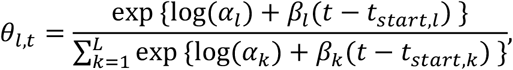

where *α*_*l*_ is the initial proportion of lineage *l*; *β*_*l*_ is a constant relative fitness growth rate of lineage *l* relative to the oldest ancestral group (Group 1); *t*_*start,l*_ is the time of emergence for lineage *l*. We used a Laplace distribution with location 0 and scale 1 as the prior for relative fitness growth rates *β*_*l*_.

For lineages present at the initial time point of the analysed time series (*t*_1_), denoted by the set *A*, their initial proportions {*α*_*l*_: *l* ∈ *A*} were modelled as a simplex vector, representing the lineage proportions at time *t*_1_ . For these lineages, the starting time was considered as *t*_*start,l*_ = *t*_1_·

For lineages that emerged after *t*_1_ (*l* ∉ *A*), their initial lineage proportion *α*_*l*_ was assigned a strong prior concentrated at small values close to zero, Beta(1,999), according to the assumption that newly emerging lineages begin at a low frequency. The time of emergence for these newly emerging lineages was defined as:

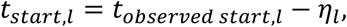

where *t*_*observed start,l*_ refers to the earliest time of nodes in lineage *l, η*_*l*_ > 0 represents the latent time between lineage origin and the first observation. To preserve phylogenetic consistency, we imposed the constraint *t*_*start,l*_ > *t*_*start,parent*(*l*)_ to ensure that descendant lineages could not originate before their parental lineage. A strongly informative prior was placed on *η*_*l*_ to constrain plausible emergence times (Supplementary Text S1).

The model was implemented in Stan *(22)*. We ran three Hamiltonian Monte Carlo (HMC) chains with 3,000 iterations each, and removed the first 1,000 iterations as burn-in. To assess sensitivity to the phylogenetic placement of internal nodes, we alternatively estimated lineage fitness using *y*_·,*t*_ values derived solely from the tips (Supplementary Text S2).

The fitness growth rate *β*_*l*_ was parameterized relative to the oldest ancestral lineage (Group 1). To evaluate fitness against a long-term co-circulating reference, we calculated the relative growth rate compared to Group 2 as exp{*β*_*l*_ − *β*_2_}· Furthermore, the fitness advantage relative to a lineage parental ancestor was calculated as exp{*β*_*l*_ − *β*_*parental*(*l*)_}. The real-time relative fitness of lineage *l* at time *t* (*r*_*l,t*_), compared to other co-circulating lineages, was estimated as exp{*θ*_*l,t*_*Σ*_*k*_[*θ*_*k,t*_(*β*_*l*_ − *β*_*k*_)]}.

### Lineage-defining substitution identification

Genomic changes were classified as lineage-defining if they were present in more than 70% of nodes within a given lineage but in less than 30% of nodes in its parental lineage. We identified lineage-defining mutations for both amino acid sites and UTR nucleotides. Internal node states were reconstructed by ancestral state reconstruction using the ape package, based on sequences with nucleotide length greater than 10,000. To evaluate the sensitivity of the results to uncertainty in ancestral state reconstruction, we conducted the analyses using terminal tips alone and incorporating both internal nodes and terminal tips. Both methods identified the same lineage-defining amino acid substitutions except at residue E473. This site was identified only in the tips-only analysis under the selected threshold because of a complex substitution pattern (V to M, with a minority of L) in Group 2 (Supplementary Fig. S2). For UTR substitutions, analyses were conducted using terminal tips alone because the higher proportion of missing data may limit the reliability of ancestral state reconstruction. The identified UTR substitutions each had an approximate missingness of 30% (Supplementary Fig. S3).

## Data Availability

All data used in this study are available at https://github.com/chenyn226/zika-phylowave.

## List of Supplementary Materials

Figure S1-S5

Text S1-S2

## Acknowledgment. Funding

This study is supported by the Wellcome Trust (Grant No. 315770/Z/24/Z to H.S. and BUZZ) and the National University of Singapore Start-Up Grant (H.C.).

## Author contributions

Conceptualization: N.L., H.S., BUZZ study team; Methodology: Y.C., D.F., N.L., H.S.; Software: Y.C., D.F.; Validation: Y.C.; Formal analysis: Y.C.; Investigation: H.S.; Resources: H.C., H.S.; Data Curation: Y.C.; Visualization: Y.C.; Supervision: H.S.; Project administration: H.S.; Funding acquisition: H.C., H.S.; Writing - Original Draft: Y.C., H.S.; Writing - Review & Editing: Y.C., D.F., H.C., N.L., H.S., the BUZZ study team.

