## Supplementary materials for "Uncovering the fitness of endemically circulating Zika virus strains"

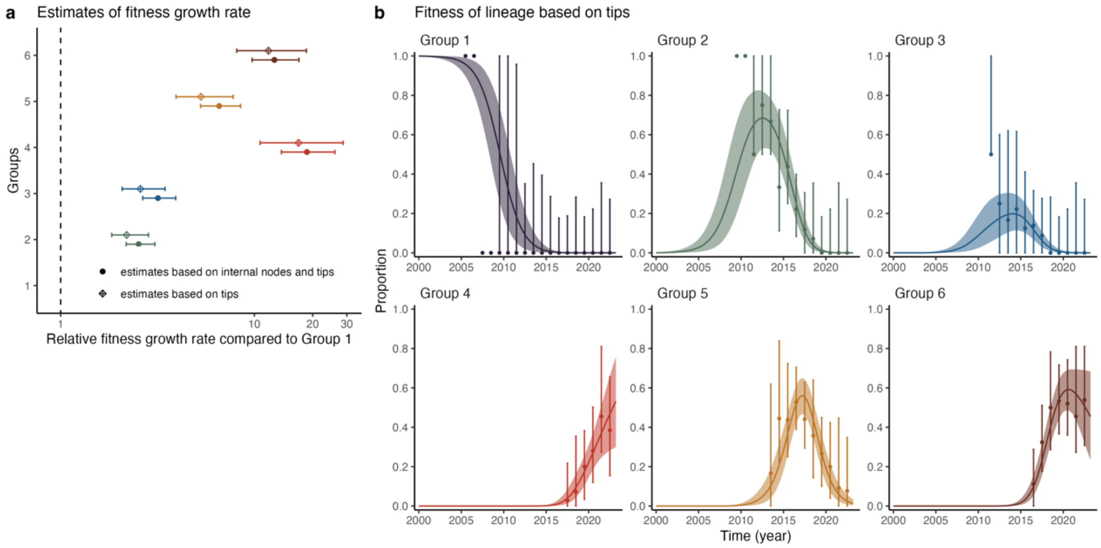


Figure S1. The fitness estimates based on lineage proportions of tips. a. The estimates of relative fitness growth rate compared to Group 1. The points and bars indicate posterior median and 95% credible intervals. b. The fitness dynamics of the identified lineages based on proportions of tips. The points and vertical bars refer to empirical values and 95% confidence intervals derived from the phylogenetic tree. The solid lines and shaded regions refer to the posterior median and 95% credible intervals.

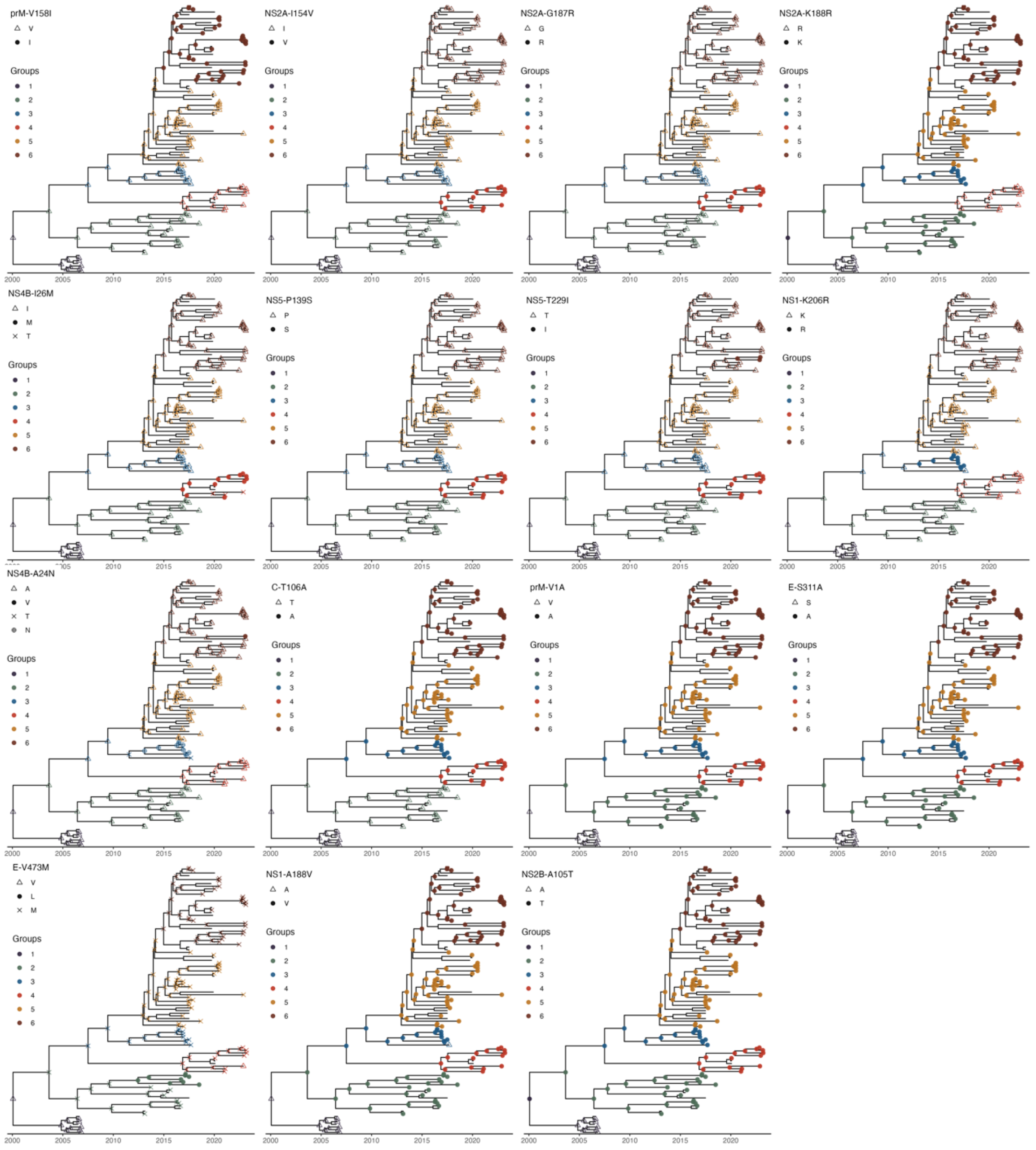


Figure S2. The distribution of lineage-defining amino acid substitutions on the phylogenetic tree.

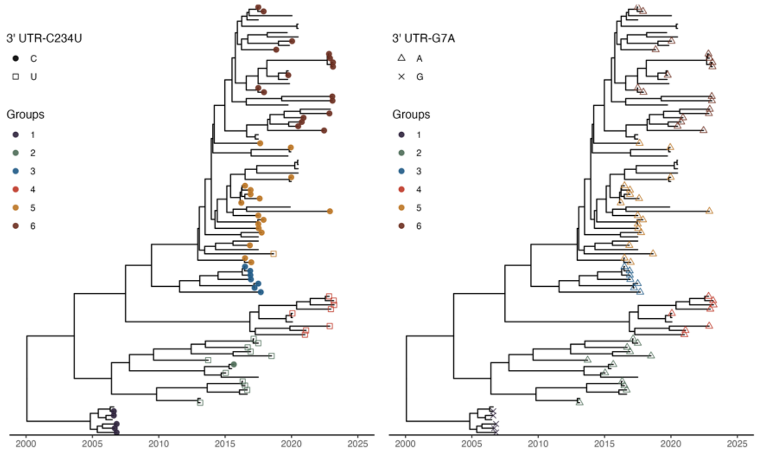


Figure S3. The distribution of lineage-defining 3' untranslated region (UTR) nucleotide substitutions on the phylogenetic tree.

| Amino acid | Residue position | Group 1 | 2 | 3 | 4 | 5 | 6 | Brazil | FP | Asian (1966) | Afican |
|---|---|---|---|---|---|---|---|---|---|---|---|
| prM | 158 | V | V | V | V | V | I | V | V | V | V |
| NS2A | 154 | I | I | I | V | I | I | I | I | I | I |
| NS2A | 187 | G | G | G | R | G | G | G | G | G | G |
| NS2A | 188 | R | K | K | R | K | K | K | K | K | K |
| NS4B | 26 | I | I | I | M | I | I | I | I | M | M |
| NS5 | 139 | P | P | P | S | P | P | P | P | S | P |
| NS5 | 229 | T | T | T | I | T | T | T | T | I | I |
| NS1 | 206 | K | K | R | K | K | K | K | K | K | K |
| NS4B | 24 | A | A | N | A | A | A | A | A | A | A |
| C | 106 | T | T | A | A | A | A | A | A | A | A |
| prM | 1 | V | A | A | A | A | A | A | A | V | A |
| E | 311 | S | A | A | A | A | A | A | A | A | T |
| E | 473 | V | M | M | M | M | M | M | M | V | L |
| NS1 | 188 | A | V | V | V | V | V | V | V | A | V |
| NS2B | 105 | A | T | T | T | T | T | T | T | T | A |
| prM | 17 | S | S | S | S | S | S | N | N | S | S |
| NS5 | 114 | M | M | M | M | M | M | V | M | T | M |

Figure S4. Lineage-defining amino acid substitutions. Rows correspond to specific residue positions, with letters representing the consensus residue for each lineage. Shaded cells indicate the presence of substitutions, with colors indicating the lineage defining each substitution. Asian (1966) refers to the early Asian lineage ZIKV detected in Malaysia in 1966 (P6-740 strain). African refers to the African lineage collected in Uganda (MR-766 strain; sequence number: NC_012532.1). Abbreviations: FP, French Polynesia.

### Materials and Methods

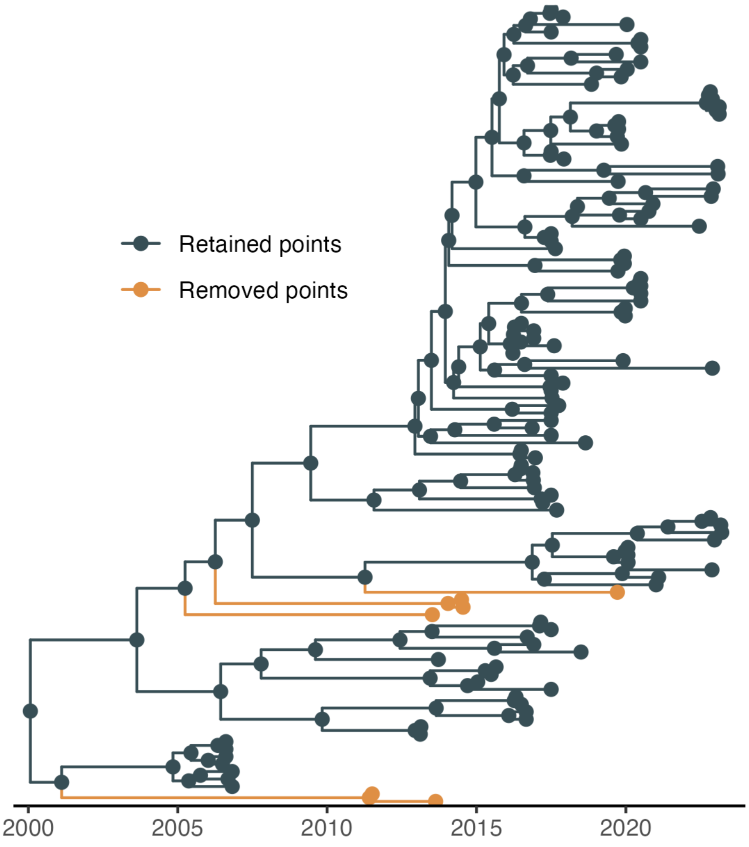


Figure S5. The sequences removed from the analysis because of the long branches.

Text S1: The prior distribution of $\eta_l$ in $t_{start,l} = t_{observed\ start,l} - \eta_l$

We constructed $\eta_l$ as $\eta_l = \eta_l^* + \rho_l \eta_{parent(l)}^*$ where $\rho_l \in [0,1]$; $\eta_l^* \in [0, c_l]$ with $c_l = \min\{c, t_{observed\ start,l} - t_{observed\ start,parent(l)}\}$ and $c$ selected as 5 years in our model. In our case, where $t_{observed\ start,l} \geq t_{observed\ start,parent(l)}$, this construction satisfies the constraint $t_{start,l} \geq t_{start,parent(l)}$. We used a strong informative prior for $\eta_l^*$ as normal distribution with mean 3 years and standard deviation 1 year, and a strong informative prior for $\rho_l$ as Beta(1,9).

Text S2: Fitness estimation model using values derived solely from the tips

Let $L$ denote the number of lineages, $y_{\cdot,t} = \{y_{l,t}: l = 1, \ldots, L\}$ denote the vector of isolate counts for each lineage $l$ at time $t$. The model using $y_{\cdot,t}$ constructed from tips is generally the same as that using $y_{\cdot,t}$ constructed from both internal nodes and tips, except for the following components. Let $t_{observed\ start\ of\ tips,l}$ refer to the earliest time of tips in lineage $l$. $y_{l,t}$ was treated as missing for $t < t_{observed\ start\ of\ tips,l}$, where $M_t$ denotes the set of lineages with missing $y_{l,t}$ at time $t$. For each time point $t$, $y_{\cdot,t}$ follows Multinomial($\theta_{\cdot,t}$) if $y_{l,t}$ is not missing for all lineages ($M_t = \emptyset$); $y_{l,t}$ follows Binomial($\Sigma_{k \notin M_t} y_{k,t}, \theta_{l,t}$) if $M_t \neq \emptyset$ and $l \notin M_t$.
